# Novel Liver Injury Phenotypes and Outcomes in Pulmonary Arterial Hypertension

**DOI:** 10.1101/2023.09.28.23296316

**Authors:** Jacqueline V Scott, Jude Moutchia, Robin L McClelland, Nadine Al-Naamani, Ethan Weinberg, Harold I Palevsky, Jasleen Minhas, Dina K Appleby, Akaya Smith, Steven C Pugliese, Corey E Ventetuolo, Steven M Kawut

## Abstract

**Background:** Pulmonary arterial hypertension (PAH) and chronic thromboembolic pulmonary hypertension (CTEPH) are disorders of the pulmonary vasculature that cause right ventricular dysfunction. Systemic consequences of right ventricular dysfunction include damage to other solid organs, such as the liver. However, the profiles and consequences of hepatic injury due to PAH and CTEPH have not been well-studied.

**Methods:** We aimed to identify underlying patterns of liver injury in a cohort of PAH and CTEPH patients enrolled in 15 randomized clinical trials conducted between 1998 and 2012. We used unsupervised machine learning to identify liver injury clusters in 13 trials and validated the findings in two additional trials. We then determined whether these liver injury clusters were associated with clinical outcomes or treatment effect heterogeneity.

**Results:** Our training dataset included 4,219 patients and our validation dataset included 1,756 patients with complete liver laboratory panels (serum total bilirubin, alkaline phosphatase, aspartate aminotransferase, alanine aminotransferase, and albumin). Using k-means clustering paired with factor analysis, we identified four unique liver phenotypes (no liver injury, hepatocellular injury, cholestatic injury, and combined injury patterns). Patients in the cholestatic injury liver cluster had the shortest time to clinical worsening and highest chance of worsening World Health Organization functional class. Randomization to the experimental arm was associated with a transition to healthier liver clusters compared to randomization to the control arm. The cholestatic injury group experienced the greatest placebo-corrected treatment benefit in terms of six-minute walk distance.

**Conclusions:** Liver injury patterns were associated with adverse outcomes in patients with PAH and CTEPH. Randomization to active treatment of pulmonary hypertension in these clinical trials had beneficial effects on liver health compared to placebo. The independent role of liver disease (often subclinical) in determining outcomes warrants prospective studies of the clinical utility of liver phenotyping for PAH prognosis and contribution to clinical disease.

## Introduction

Pulmonary arterial hypertension (PAH) and chronic thromboembolic pulmonary hypertension (CTEPH) are disorders of the pulmonary vasculature where elevations in pulmonary vascular resistance and mean pulmonary arterial pressure cause right ventricular dysfunction and eventual failure.^1^ PAH is being increasingly recognized as a systemic disease that involves other organs. Although it is well-established that right ventricular adaptation is the primary determinant of outcomes in PAH, the hepatic consequences of right ventricular dysfunction have not been well-studied. Specifically, right ventricular dysfunction leads to venous congestion and reduced perfusion, both of which can cause subclinical, acute, and/or chronic solid organ injury.^2^ Studies of congestive heart failure suggest that venous congestion leads to congestive hepatopathy, characterized by elevations in total bilirubin, alkaline phosphatase, and gamma glutamate transferase. Chronic venous congestion is also commonly seen in pulmonary hypertension, which can lead to cholestatic liver injury.^3^ Further, cholestatic liver injury causes impaired synthetic liver function and abnormal hepatic drug metabolism, both of which could affect outcomes in pulmonary hypertension. While the 2022 European Respiratory Society guidelines propose routine laboratory testing for liver function in patients with PAH^1^, to our knowledge there are no established clinical or subclinical profiles of liver injury or dysfunction or associations with outcome (except total bilirubin) for PAH patients.

We used unsupervised machine learning to determine underlying patterns in common liver test values in patients recruited into randomized clinical trials of PAH and CTEPH therapies submitted to the FDA. We hypothesized that certain phenotypes of liver disease defined by liver test patterns would be associated with reduced functional capacity, quality of life, time to clinical worsening, and survival. Further, we hypothesized that treatment could affect liver health and that there would be heterogeneity of treatment effects amongst baseline liver phenotypes.

## Methods

### Study and Participant Selection

The U.S. Food and Drug Administration (FDA) provided individual participant data (IPD) from 21 randomized clinical trials (RCTs) of pulmonary hypertension therapies conducted between 1998 and 2014 (Table S1). We included phase III RCTs and limited our analysis to adult patients with PAH or CTEPH. We included participants with complete data for the following liver tests at baseline: serum total bilirubin (TB), alkaline phosphatase (AlkP), aspartate aminotransferase (AST), alanine aminotransferase (ALT), and albumin. We excluded RCTs that were not submitted to the FDA and RCTs of sitaxentan, which directly caused idiosyncratic severe liver injury.^4^ Virtually all/all clinical trials excluded patients with portopulmonary hypertension, however a small number were included (n = 13, 0.3 %). All trials were performed under the supervision of human ethics committees, and all patients provided informed consent to participate in the trials. Harmonization and analysis of these data were exempted from review by the University of Pennsylvania Institutional Review Board.

## Data and Harmonization

Individual demographic and clinical data, including subject start date, anthropometrics, pulmonary hypertension etiology, six-minute walk distance (6MWD), laboratory values, and invasive hemodynamics, were harmonized across all studies using the Study Data Tabulation Model version 1.4, as described in the Study Data Tabulation Model Implementation Guide: Human Clinical Trials, Version 3.2 and as previously published.^5,6^ We calculated the Model for End-stage Liver Disease-sodium score.^7,8^ Invasive hemodynamics were captured if they were measured within 90 days of study reference start date. As the trials occurred over a period of 16 years (1998–2014), pulmonary hypertension etiology was characterized by the 2019 World Symposium on Pulmonary Hypertension classification. For studies adhering to Study Data Tabulation Model guidelines, the subject reference start date (corresponding to the date on which each subject was first administered study treatment) was most consistently recorded and used as the start date. If this was not available, the date of subject randomization was used instead.

Clinical worsening was harmonized across trials and defined as all-cause death, lung transplantation, atrial septostomy, hospitalization for worsening PAH, discontinuation of study treatment for worsening PAH, initiation of parenteral prostacyclin analogue therapy, or a decrease of at least 15% in six minute walk distance from baseline with either (i) worsening of WHO functional class from baseline or (ii) the addition of approved PAH treatment (endothelin receptor antagonists, phosphodiesterase-5 inhibitors, inhaled or oral prostacyclin analogues).

## Statistical Analysis

We split the data into a training set (approximately 70% of subjects, older trials that ended in 2012) and a validation set (approximately 30% of subjects, more recent trials that ended in 2014) to ensure sufficient sample size and generalizability of the clustering results to present-day patients. For feature selection, we performed factor analysis with varimax rotation using baseline liver biomarkers. We then performed K-means clustering on data transformed to the factor dimensional space and selected the number of clusters through the elbow method, a standard technique for optimizing the number of clusters without becoming overly granular.^9,10^ We derived K-means clusters in the training set and independently re-derived the K-means cluster centers in the validation set. We assessed whether the liver profiles in the clusters in the training set were similar to those in the re-derived clusters in the validation data set.

We assessed the comparability of demographic and clinical profiles between included and excluded patients in the training and validation sets using standardized differences. We compared demographic and clinical profiles across clusters in the derivation and validation datasets using chi-squared tests for categorical variables and one-way analysis of variance or Kruskal-Wallis tests for continuous variables.

For the longitudinal analyses, we assigned patients in the validation set and follow-up liver biomarker data for the entire cohort to clusters using the K-means cluster centers from the training set. Longitudinal changes in cluster assignment were evaluated for subjects from baseline to week 12 or 16 (whichever was available). Patients with liver biomarkers assessed at both weeks 12 and 16 had these values averaged for the analysis. If patients were missing liver biomarkers at both week 12 and 16, data at week 4 or week 8 were used. Association between treatment arm assignment and cluster assignment at week 12 or 16 was assessed using multilevel ordinal regression (cumulative link model), where each phenotype was ordered from best liver function to worst liver function as determined by clinical profile. This model accounted for baseline cluster assignment, treatment arm assignment, age, sex, baseline 6MWD, pulmonary hypertension etiology, and body mass index as fixed effects and study as a random effect. All analyses considered combination therapy as the “active” arm and monotherapy arms combined as the “control” arm for AMBITION, which compared taladafil and ambrisentan combination therapy to treatment with tadalafil or ambrisentan alone as monotherapy.

We assessed the association between baseline liver cluster and outcomes using one-step multilevel models. We used Kaplan-Meier curves and Cox proportional hazards regression with a frailty term for time to clinical worsening, random effects linear regression for change in 6MWD and quality of life (assessed by SF-36 physical component score [PCS] and mental component score [MCS]) at 12 or 16 weeks, and multilevel ordinal regression (cumulative link model) adjusted for baseline value for WHO functional class at 12 or 16 weeks. These models were adjusted for age, sex, body mass index, pulmonary hypertension etiology, and treatment arm assignment defined *a priori*. In the subset of studies with hemodynamic data within 90 days before the reference start date, we additionally adjusted for time-varying right atrial pressure and cardiac index.

We assessed effect modification of treatment arm assignment by baseline liver injury phenotype (treatment–liver injury phenotype interaction) for each outcome by including an interaction term in one-step models while separating within– and across-trial interaction.^11^

## Results

### Study population and study sample

Of the 21 available trials, we included 15 Phase III trials (13 Phase III trials in the training set and 2 Phase III trials in the validation set) in this analysis (Figure 1). Table S1 shows the trials considered for inclusion along with any pertinent liver function exclusion criteria. Of the included trials, 11 trials captured invasive hemodynamics at baseline (excludes TRIUMPH, FREEDOM-C, FREEDOM-M, FREEDOM-C2). From 6429 subjects in 16 trials, we excluded 57 (0.8%) patients aged less than 18 years and 397 (6%) adult patients missing at least one liver test at baseline (203 (51%) of which were from AIR, which did not have liver test data), leaving 5,975 patients in the study sample (4,219 patients in the training set and 1,756 patients in the validation set).

**Figure 1:**
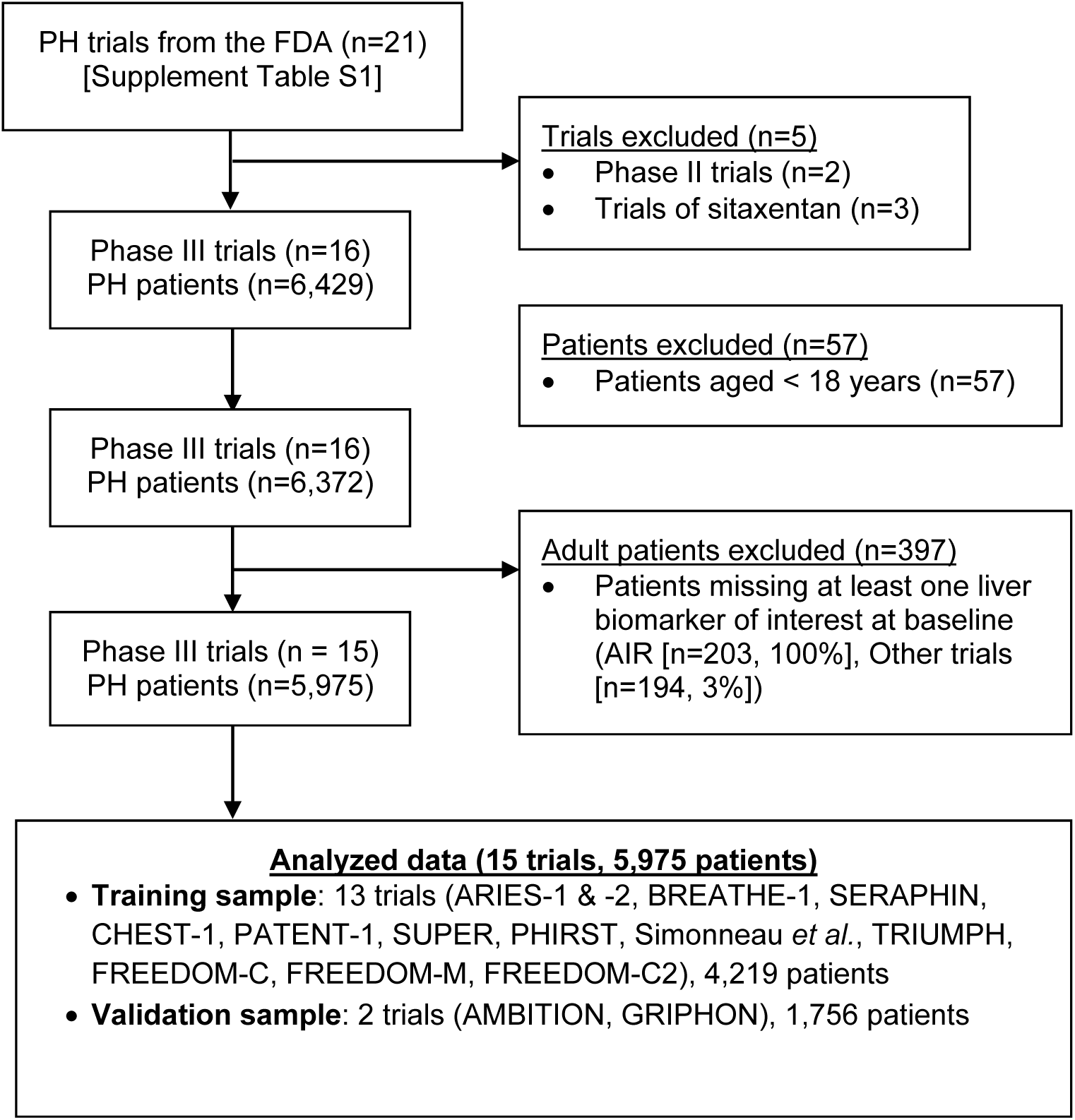
Participant flowchart.

The baseline demographic and clinical profiles of included and excluded adult pulmonary hypertension patients were mostly similar (with differences generally related to AIR II subjects being excluded due to lack of availability of liver tests) (Table S2). Included participants may have had a higher proportion of connective tissue disease and less CTEPH than the excluded patients. The study sample had more WHO functional class II participants compared to excluded patients, who had more functional class IV patients. Cardiac index was higher and pulmonary vascular resistance lower in the study sample.

The mean age of the training sample was 49.7 ± 15.1 years; 77.7% were female, and 58.0% had idiopathic PAH (Table S3). The mean age of the validation sample was 50.9 ± 15.5 years; 78.6% were female, and 55.6% had idiopathic PAH. The baseline demographic and clinical profiles of the training and validation sets were comparable.

## K-means clustering

Results from the K-means cluster analysis in the derivation sample are shown within the three-dimensional factor space in Figure 2. Plots show each cluster as it would be oriented in the factor dimensional space. Results of the factor analysis in the derivation sample with varimax rotation are shown in Figure S1. There were three distinct factors identified among the five liver tests: Factor 1) alanine aminotransferase and aspartate aminotransferase, considered measures of hepatocellular injury, 2) total bilirubin and alkaline phosphatase, considered measures of cholestatic injury, and 3) albumin, reflecting synthetic liver function. Using the three factors, 4-5 clusters were found to be optimal in K-means clustering (through the elbow method, clusters naturally occur on a sliding scale of each of these factors). Cluster 1 had the most normal values (lowest aminotransferases, lowest total bilirubin and alkaline phosphatase, highest albumin) and the phenotype was termed “no liver injury” (Table S4). Cluster 2 only had elevations in aminotransferases, consistent with hepatocellular injury and the phenotype was termed “hepatocellular injury pattern”. Cluster 3 had higher total bilirubin and alkaline phosphatase values (consistent with a cholestatic pattern) and lower albumin levels and the phenotype was termed “cholestatic injury pattern”. Clusters 4 and 5 had the most abnormal values for all tests and were combined due to their proximity in the factor dimensional space and only having 5 individuals in Cluster 5 and was termed “combined injury pattern”.

**Figure 2.**
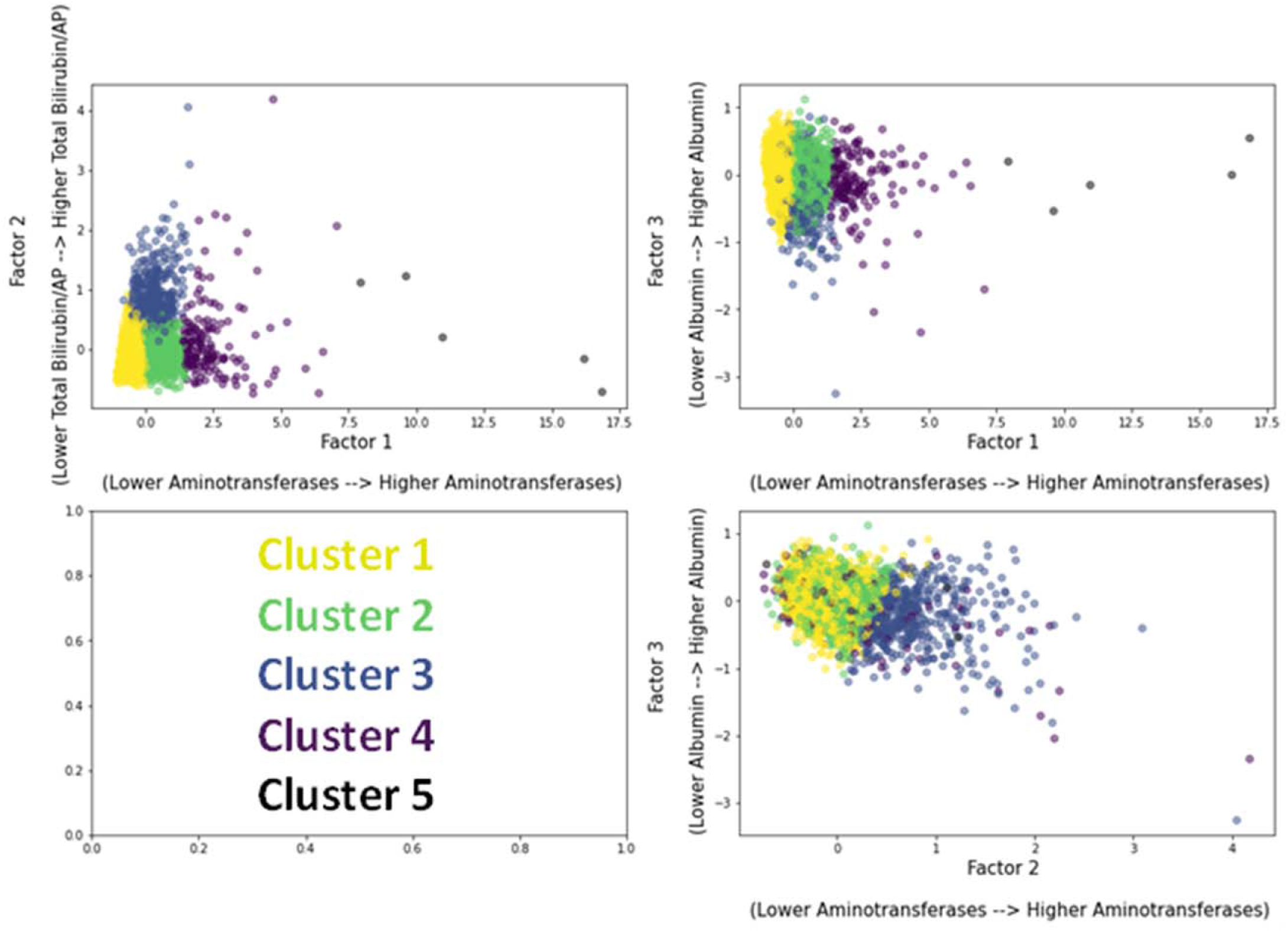
K-means clustering results in factor dimensional space.

The four phenotypes had similar distributions of age and body mass index, some comorbidities including diabetes mellitus and hypertension, and treatment arm assignment (Table 1). The “no liver injury” phenotype had the highest proportion of females, the lowest prevalence of pre-existing liver disease, lowest NTproBNP and BNP values, and the least affected baseline hemodynamics (lowest mean right atrial pressure, mean pulmonary arterial pressure, and pulmonary vascular resistance, and highest cardiac index). Compared to the “no liver injury” phenotype, the “hepatocellular injury pattern” phenotype had higher NTproBNP and BNP levels, and slightly worse hemodynamics. The “cholestatic injury pattern” phenotype had the lowest proportion of female patients, a higher prevalence of preexisting liver disease, the highest NTproBNP and BNP values, and the worst hemodynamics, including both lower cardiac index and higher mean right atrial pressure, consistent with more venous congestion. The “cholestatic injury pattern” phenotype also had the lowest 6MWD and the highest MELD-Na score.

**Table 1:**
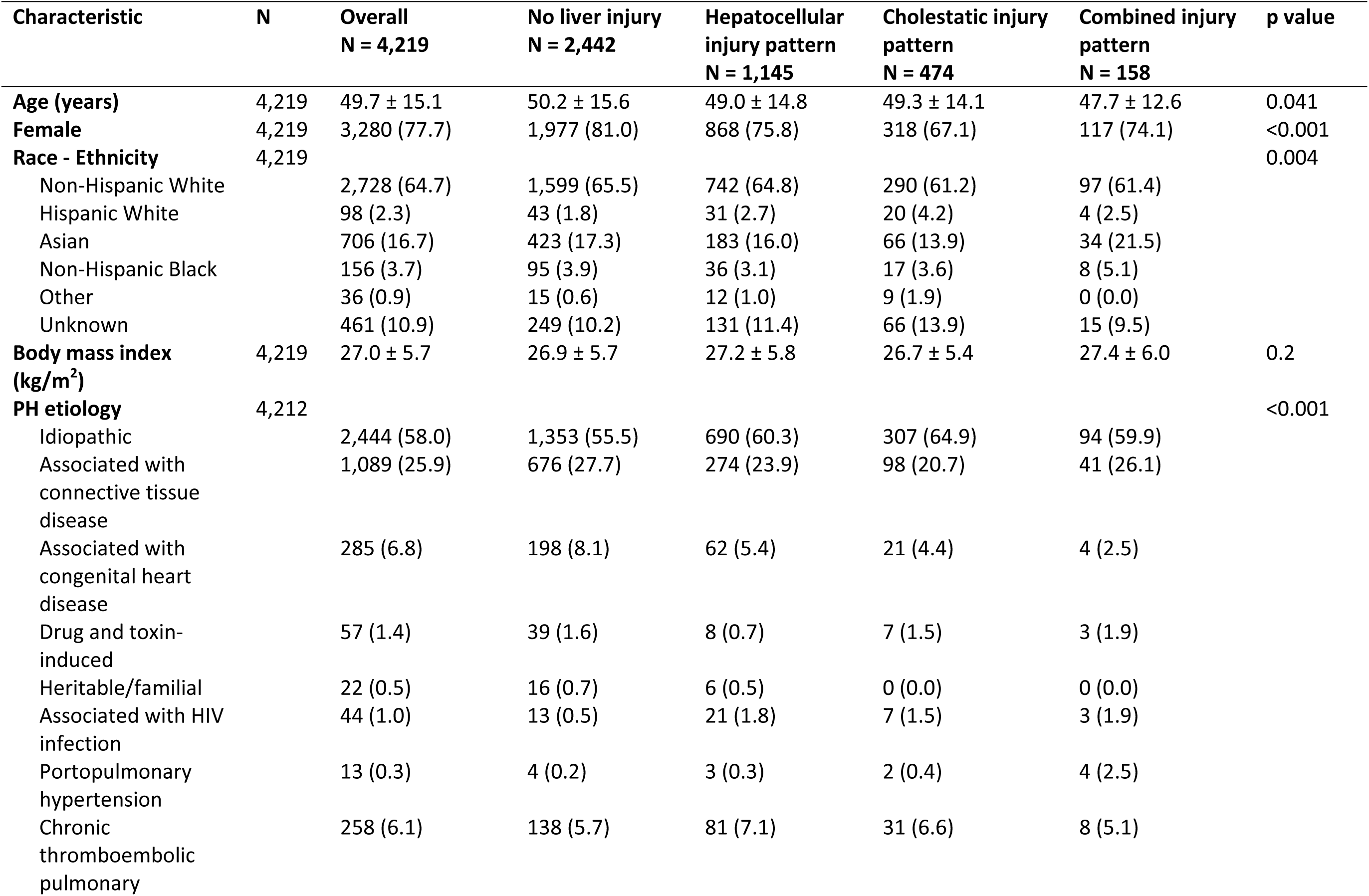
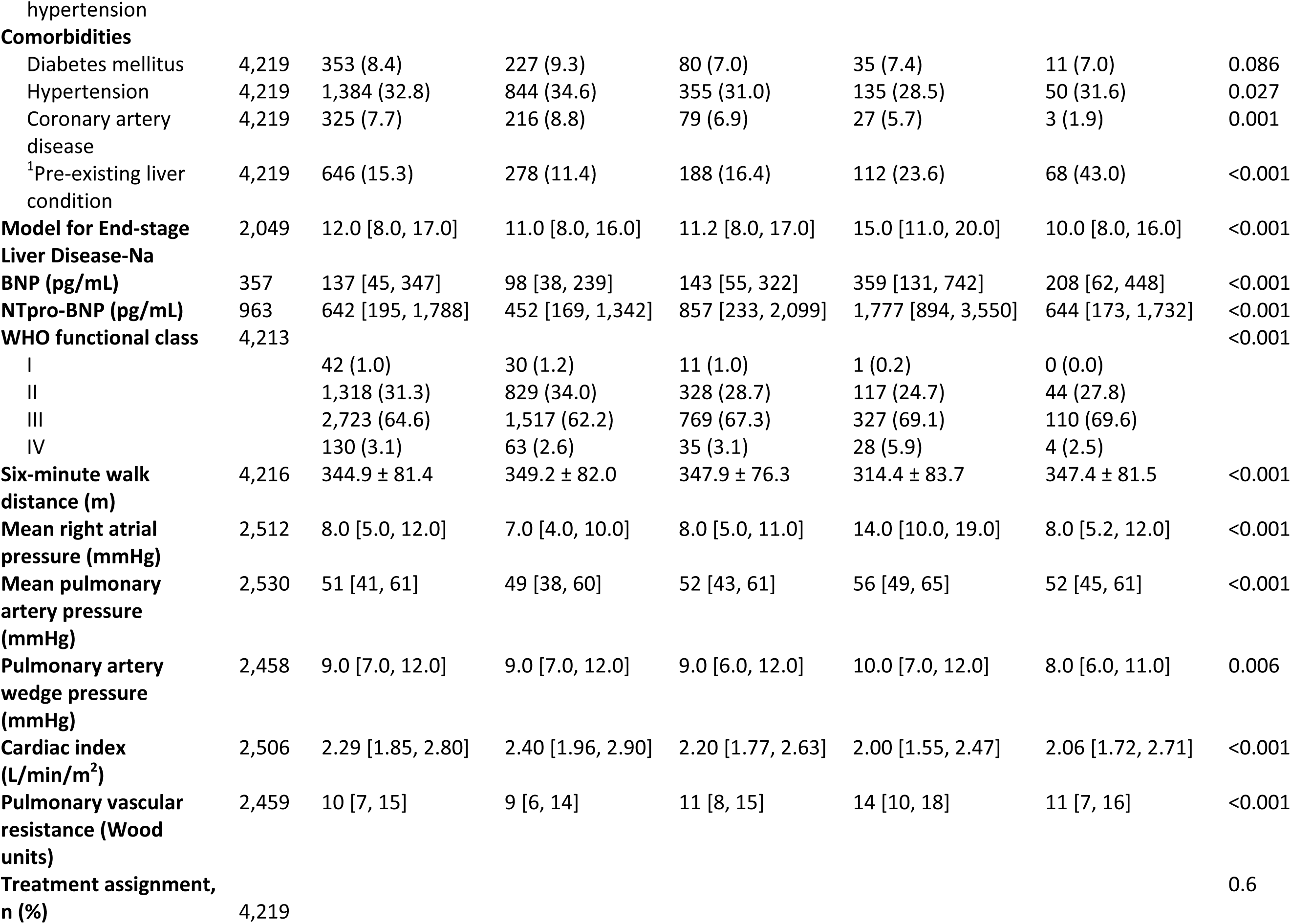
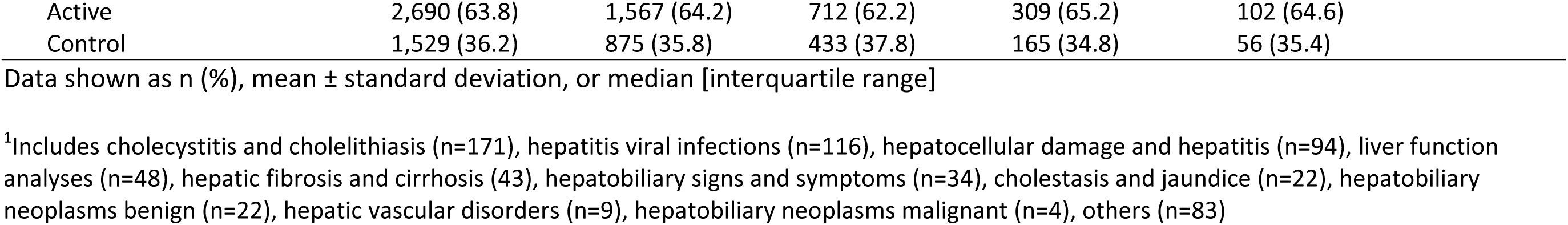
Baseline demographic and clinical data by liver injury phenotype (Training Data)

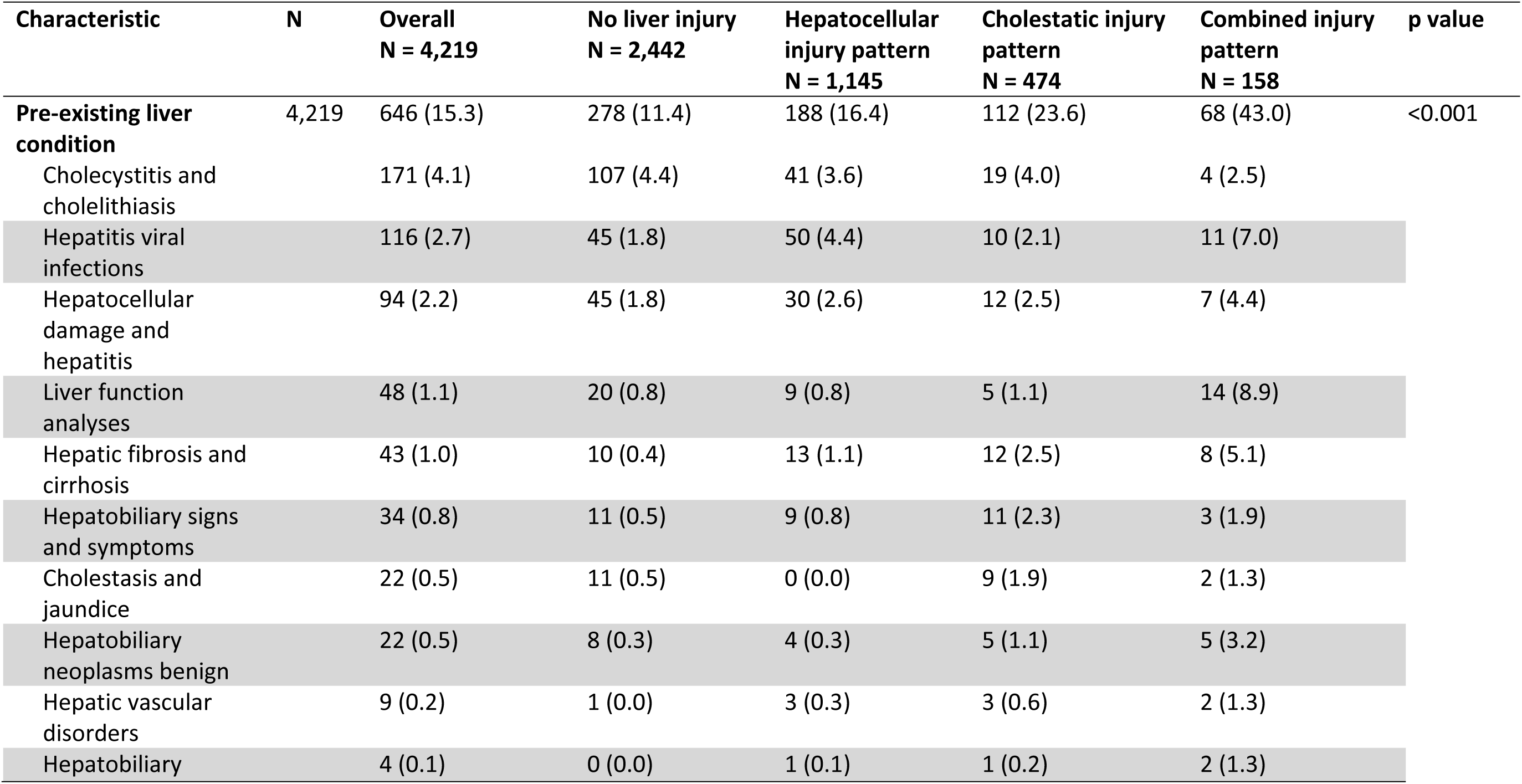
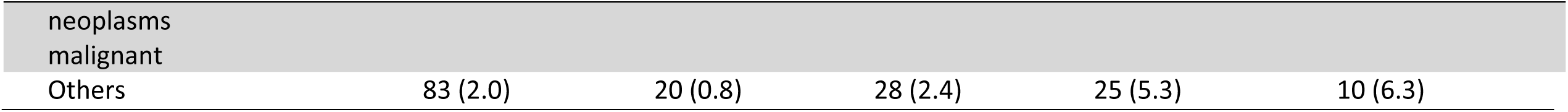

| Characteristic | N | Overall<br>N = 4,219 | No liver injury<br>N = 2,442 | Hepatocellular<br>injury pattern<br>N = 1,145 | Cholestatic injury<br>pattern<br>N = 474 | Combined injury<br>pattern<br>N = 158 | p value |
| --- | --- | --- | --- | --- | --- | --- | --- |
| **Pre-existing liver condition** | 4,219 | 646 (15.3) | 278 (11.4) | 188 (16.4) | 112 (23.6) | 68 (43.0) | <0.001 |
| Cholecystitis and cholelithiasis |  | 171 (4.1) | 107 (4.4) | 41 (3.6) | 19 (4.0) | 4 (2.5) |  |
| Hepatitis viral infections |  | 116 (2.7) | 45 (1.8) | 50 (4.4) | 10 (2.1) | 11 (7.0) |  |
| Hepatocellular damage and hepatitis |  | 94 (2.2) | 45 (1.8) | 30 (2.6) | 12 (2.5) | 7 (4.4) |  |
| Liver function analyses |  | 48 (1.1) | 20 (0.8) | 9 (0.8) | 5 (1.1) | 14 (8.9) |  |
| Hepatic fibrosis and cirrhosis |  | 43 (1.0) | 10 (0.4) | 13 (1.1) | 12 (2.5) | 8 (5.1) |  |
| Hepatobiliary signs and symptoms |  | 34 (0.8) | 11 (0.5) | 9 (0.8) | 11 (2.3) | 3 (1.9) |  |
| Cholestasis and jaundice |  | 22 (0.5) | 11 (0.5) | 0 (0.0) | 9 (1.9) | 2 (1.3) |  |
| Hepatobiliary neoplasms benign |  | 22 (0.5) | 8 (0.3) | 4 (0.3) | 5 (1.1) | 5 (3.2) |  |
| Hepatic vascular disorders |  | 9 (0.2) | 1 (0.0) | 3 (0.3) | 3 (0.6) | 2 (1.3) |  |
| Hepatobiliary |  | 4 (0.1) | 0 (0.0) | 1 (0.1) | 1 (0.2) | 2 (1.3) |  |

|  |  |  |  |  |  |
| --- | --- | --- | --- | --- | --- |
| neoplasms<br>malignant |  |  |  |  |  |
| Others | 83 (2.0) | 20 (0.8) | 28 (2.4) | 25 (5.3) | 10 (6.3) |

Similar clusters were identified in the validation dataset with the same liver function test patterns as in the training set (Table S4). In the validation dataset, Cluster 1 included the healthiest patients (in terms of liver test pattern), cluster 2 showed higher aminotransferases (hepatocellular injury pattern), cluster 3 showed higher bilirubin and alkaline phosphatase (cholestatic pattern), and clusters 4 and 5 also had elevations for all liver tests. The distribution of baseline demographic and clinical data across phenotypes in the validation dataset was similar to that observed in the training dataset (Table 2). Due to the similarities between the training and validation clusters, we assigned patients in the validation set and with follow-up liver test data for the entire cohort to clusters using the K-means cluster centers from the training set.

**Table 2:** Baseline demographic and clinical profile by liver injury phenotype (Validation Data)

| Characteristic | N | Overall<br>N = 1,756 | No liver injury<br>N = 1,112 | Hepatocellular<br>injury pattern<br>N = 386 | Cholestatic injury<br>pattern<br>N = 200 | Combined injury<br>pattern<br>N = 58 | p value |
| --- | --- | --- | --- | --- | --- | --- | --- |
| **Age (years)** | 1,756 | 50.9 ± 15.5 | 51.0 ± 16.0 | 50.9 ± 14.6 | 50.4 ± 14.2 | 50.4 ± 15.4 | 0.9 |
| **Female** | 1,756 | 1,381 (78.6) | 914 (82.2) | 280 (72.5) | 142 (71.0) | 45 (77.6) | <0.001 |
| **Race - Ethnicity** | 1,756 |  |  |  |  |  | 0.029 |
| Non-Hispanic White |  | 1,236 (70.4) | 792 (71.2) | 266 (68.9) | 139 (69.5) | 39 (67.2) |  |
| Hispanic White |  | 54 (3.1) | 36 (3.2) | 10 (2.6) | 4 (2.0) | 4 (6.9) |  |
| Asian |  | 257 (14.6) | 162 (14.6) | 54 (14.0) | 30 (15.0) | 11 (19.0) |  |
| Non-Hispanic Black |  | 65 (3.7) | 47 (4.2) | 14 (3.6) | 2 (1.0) | 2 (3.4) |  |
| Hispanic Black |  |  |  |  |  |  |  |
| Other |  | 1 (0.1) | 0 (0.0) | 1 (0.3) | 0 (0.0) | 0 (0.0) |  |
| Unknown |  | 26 (1.5) | 15 (1.3) | 9 (2.3) | 1 (0.5) | 1 (1.7) |  |
| **Body mass index**<br>**(kg/m<sup>2</sup>)** | 1,756 | 27.5 ± 5.8 | 27.5 ± 5.8 | 27.7 ± 6.0 | 27.8 ± 5.6 | 26.2 ± 5.3 | 0.3 |
| **PH etiology** | 1,755 |  |  |  |  |  | <0.001 |
| Idiopathic |  | 975 (55.6) | 601 (54.0) | 221 (57.4) | 129 (64.5) | 24 (41.4) |  |
| Associated with<br>connective tissue<br>disease |  | 549 (31.3) | 349 (31.4) | 120 (31.2) | 52 (26.0) | 28 (48.3) |  |
| Associated with<br>congenital heart<br>disease |  | 118 (6.7) | 94 (8.5) | 15 (3.9) | 8 (4.0) | 1 (1.7) |  |
| Drug and toxin-<br>induced |  | 50 (2.8) | 31 (2.8) | 12 (3.1) | 4 (2.0) | 3 (5.2) |  |
| Heritable/familial |  | 43 (2.5) | 30 (2.7) | 11 (2.9) | 2 (1.0) | 0 (0.0) |  |
| Associated with HIV<br>infection |  | 20 (1.1) | 7 (0.6) | 6 (1.6) | 5 (2.5) | 2 (3.4) |  |
| **Comorbidities** |  |  |  |  |  |  |  |
| Diabetes mellitus | 1,756 | 226 (12.9) | 141 (12.7) | 46 (11.9) | 30 (15.0) | 9 (15.5) | 0.7 |
| Hypertension | 1,756 | 669 (38.1) | 421 (37.9) | 154 (39.9) | 75 (37.5) | 19 (32.8) | 0.7 |
| Coronary artery | 1,756 | 192 (10.9) | 120 (10.8) | 40 (10.4) | 26 (13.0) | 6 (10.3) | 0.8 |
| disease |  |  |  |  |  |  |  |
| <sup>1</sup>Pre-existing liver condition | 1,756 | 276 (15.7) | 146 (13.1) | 63 (16.3) | 46 (23.0) | 21 (36.2) | <0.001 |
| **NTpro-BNP (pg/mL)** | 1,736 | 628 [208, 1,750] | 457 [163, 1,278] | 834 [303, 1,906] | 1,947 [1,032, 3,018] | 916 [346, 2,086] | <0.001 |
| **WHO functional class** | 1,756 |  |  |  |  |  | 0.1 |
| I |  | 9 (0.5) | 7 (0.6) | 1 (0.3) | 1 (0.5) | 0 (0.0) |  |
| II |  | 719 (40.9) | 478 (43.0) | 150 (38.9) | 65 (32.5) | 26 (44.8) |  |
| III |  | 1,017 (57.9) | 617 (55.5) | 235 (60.9) | 133 (66.5) | 32 (55.2) |  |
| IV |  | 11 (0.6) | 10 (0.9) | 0 (0.0) | 1 (0.5) | 0 (0.0) |  |
| **Six-minute walk distance (m)** | 1,756 | 351.0 ± 84.2 | 356.2 ± 83.3 | 353.1 ± 78.8 | 321.5 ± 91.8 | 338.2 ± 89.2 | <0.001 |
| **Mean right atrial pressure (mmHg)** | 1,547 | 8.0 [5.0, 11.0] | 7.0 [5.0, 10.0] | 7.5 [5.0, 11.0] | 11.0 [7.0, 16.0] | 8.0 [5.0, 11.0] | <0.001 |
| **Mean pulmonary artery pressure (mmHg)** | 1,702 | 50 [41, 60] | 49 [40, 58] | 53 [42, 61] | 54 [45, 62] | 51 [43, 58] | <0.001 |
| **Pulmonary artery wedge pressure (mmHg)** | 1,606 | 9.0 [7.0, 12.0] | 9.0 [7.0, 12.0] | 9.0 [7.0, 12.0] | 10.0 [7.0, 12.0] | 10.0 [6.0, 12.0] | 0.4 |
| **Cardiac index (L/min/m<sup>2</sup>)** | 1,636 | 2.40 [1.96, 2.90] | 2.48 [2.02, 2.95] | 2.23 [1.90, 2.70] | 2.12 [1.79, 2.63] | 2.40 [1.92, 2.84] | <0.001 |
| **Pulmonary vascular resistance (Wood units)** | 1,663 | 9.4 [6.7, 13.4] | 8.8 [6.3, 12.5] | 10.9 [7.7, 14.7] | 11.3 [7.8, 15.7] | 10.0 [7.7, 13.6] | <0.001 |
| **Treatment assignment, n (%)** | 1,756 |  |  |  |  |  | 0.7 |
| Active |  | 876 (49.9) | 556 (50.0) | 199 (51.6) | 93 (46.5) | 28 (48.3) |  |
| Control |  | 880 (50.1) | 556 (50.0) | 187 (48.4) | 107 (53.5) | 30 (51.7) |  |

Data shown as n (%), mean ± standard deviation, or median [interquartile range]
<sup>1</sup>Includes cholecystitis and cholelithiasis (n=87), hepatitis viral infections (n=60), hepatocellular damage and hepatitis (n=53), hepatic fibrosis and cirrhosis (22), liver function analyses (n=13), cholestasis and jaundice (n=10), hepatobiliary neoplasms benign (n=7), hepatobiliary signs and symptoms (n=6), hepatic vascular disorders (n=2), hepatobiliary neoplasms malignant (n=1), others (n=15)

| Characteristic | N | Overall<br>N = 1,756 | No liver injury<br>N = 1,112 | Hepatocellular<br>injury pattern<br>N = 386 | Cholestatic injury<br>pattern<br>N = 200 | Combined injury<br>pattern<br>N = 58 | p value |
| --- | --- | --- | --- | --- | --- | --- | --- |
| **Pre-existing liver condition** | 1,756 | 276 (15.7) | 146 (13.1) | 63 (16.3) | 46 (23.0) | 21 (36.2) | <0.001 |
| Cholecystitis and cholelithiasis |  | 87 (5.0) | 56 (5.0) | 14 (3.6) | 13 (6.5) | 4 (6.9) |  |
| Hepatitis viral infections |  | 60 (3.4) | 33 (3.0) | 15 (3.9) | 6 (3.0) | 6 (10.3) |  |
| Hepatocellular damage and hepatitis |  | 53 (3.0) | 31 (2.8) | 13 (3.4) | 4 (2.0) | 5 (8.6) |  |
| Hepatic fibrosis and cirrhosis |  | 22 (1.3) | 7 (0.6) | 7 (1.8) | 5 (2.5) | 3 (5.2) |  |
| Liver function analyses |  | 13 (0.7) | 5 (0.4) | 3 (0.8) | 4 (2.0) | 1 (1.7) |  |
| Cholestasis and jaundice |  | 10 (0.6) | 4 (0.4) | 1 (0.3) | 5 (2.5) | 0 (0.0) |  |
| Hepatobiliary neoplasms benign |  | 7 (0.4) | 4 (0.4) | 1 (0.3) | 2 (1.0) | 0 (0.0) |  |
| Hepatobiliary signs and symptoms |  | 6 (0.3) | 3 (0.3) | 1 (0.3) | 1 (0.5) | 1 (1.7) |  |
| Hepatic vascular disorders |  | 2 (0.1) | 0 (0.0) | 0 (0.0) | 1 (0.5) | 1 (1.7) |  |
| Hepatobiliary neoplasms malignant |  | 1 (0.1) | 0 (0.0) | 0 (0.0) | 1 (0.5) | 0 (0.0) |  |
| Others |  | 15 (0.9) | 3 (0.3) | 8 (2.1) | 4 (2.0) | 0 (0.0) |  |

## Longitudinal changes in liver injury phenotype by treatment assignment

A total of N = 4,932 patients had liver biomarker data available at either Week 4, 8, 12, or 16 (most patients had liver biomarkers available at week 12 and/or 16, N = 3,094, and the remaining patients, N = 1,838, only had liver biomarkers available at week 4 or 8). Most patients that were excluded from longitudinal analysis had a single value missing and therefore could not be clustered). There were differences in patients transitioning between liver injury phenotypes even during the brief interval between baseline and week 12 or 16 in the active treatment arm and control arm (Figure 3). Randomization to active treatment was associated with a significantly lower odds of transitioning to a “worse” liver injury phenotype at 12 or 16 weeks (adjusted OR: 0.67 [95% CI: 0.56, 0.81], p < 0.001) compared to randomization to control. The interaction between treatment and baseline phenotype was not significant (p = 0.62), meaning that the type of baseline liver injury pattern did not determine how much active treatment vs placebo allocation impacted on the transition to other liver injury patterns. Notable findings included more patients in the “cholestatic injury” phenotype treated with active therapy transitioning to the “no liver injury” and “hepatocellular injury” phenotypes, compared to those in the control group who were more likely to transition from the “hepatocellular injury” phenotype to the cholestatic injury phenotype and from the “no liver injury” group to the “hepatocellular injury pattern” group.

**Figure 3:**
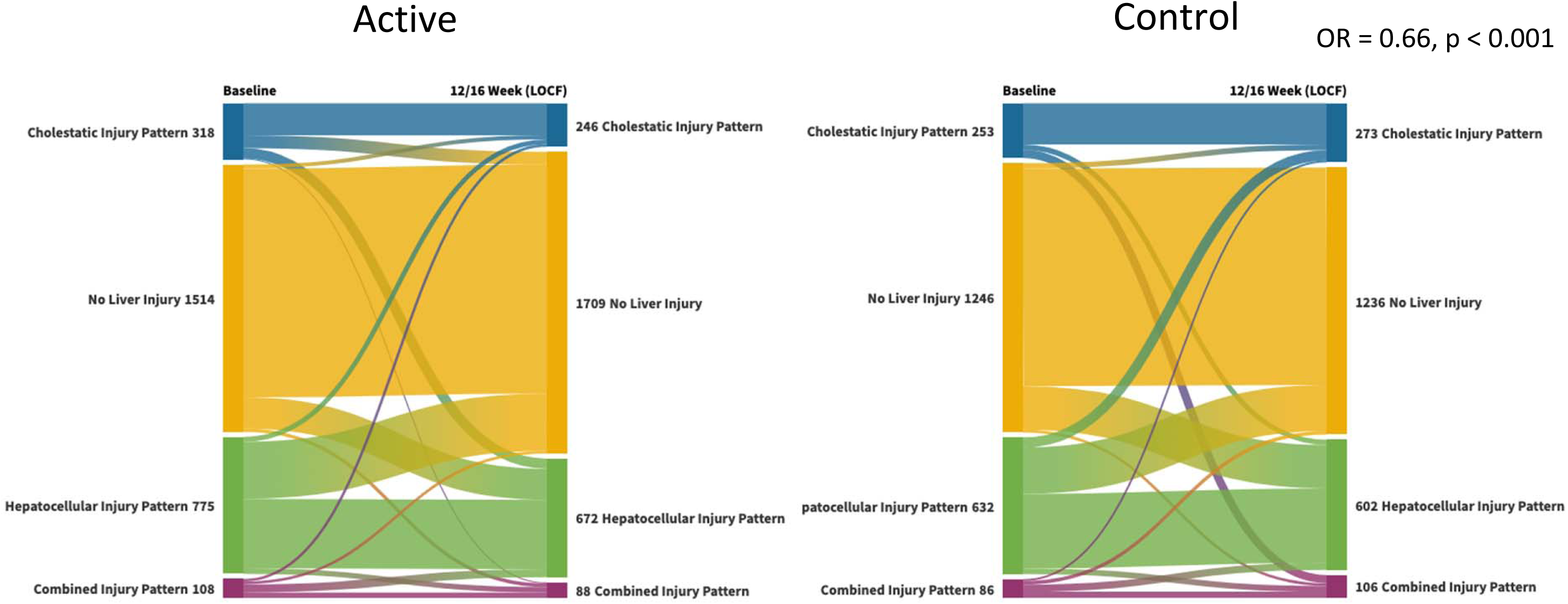
Changes in liver injury phenotype from baseline to week 12/16 for assignment to active vs control arms.

## Association between liver injury phenotype and outcomes

A total of 1,150 patients (249 per 1,000 person-years) had at least one clinical worsening event during follow-up: 675 (207 per 1,000 person-years) patients in the “no liver injury” group, 244 (288 per 1,000 person-years) in the “hepatocellular injury pattern” group, 177 (467 per 1,000 person-years) in the “cholestatic injury pattern” group, and 44 (382 per 1,000 person-years) in the “combined injury pattern” group. Time to clinical worsening stratified by liver injury phenotype at baseline is shown in Figure 4. The “hepatocellular injury pattern”, “cholestatic injury pattern”, and “combined injury pattern” phenotypes had significantly higher likelihood of having a clinical worsening event compared to the “no liver injury” phenotype after adjustment for age, sex, body mass index, etiology of pulmonary hypertension, baseline 6MWD, and treatment assignment (Table 3). These associations persisted when only death or hospitalization for worsening PH were included as outcomes and with additional adjustment for baseline and (for those who had it available) follow-up right atrial pressure and cardiac index. Also, the results were similar when liver injury phenotype was modeled as a time-varying covariate (i.e., accounting for the change in liver injury phenotype from baseline to 12/16 weeks) (Table S5). Patients with the “cholestatic injury pattern” had even more pronounced increased risk in this analysis.

**Figure 4:**
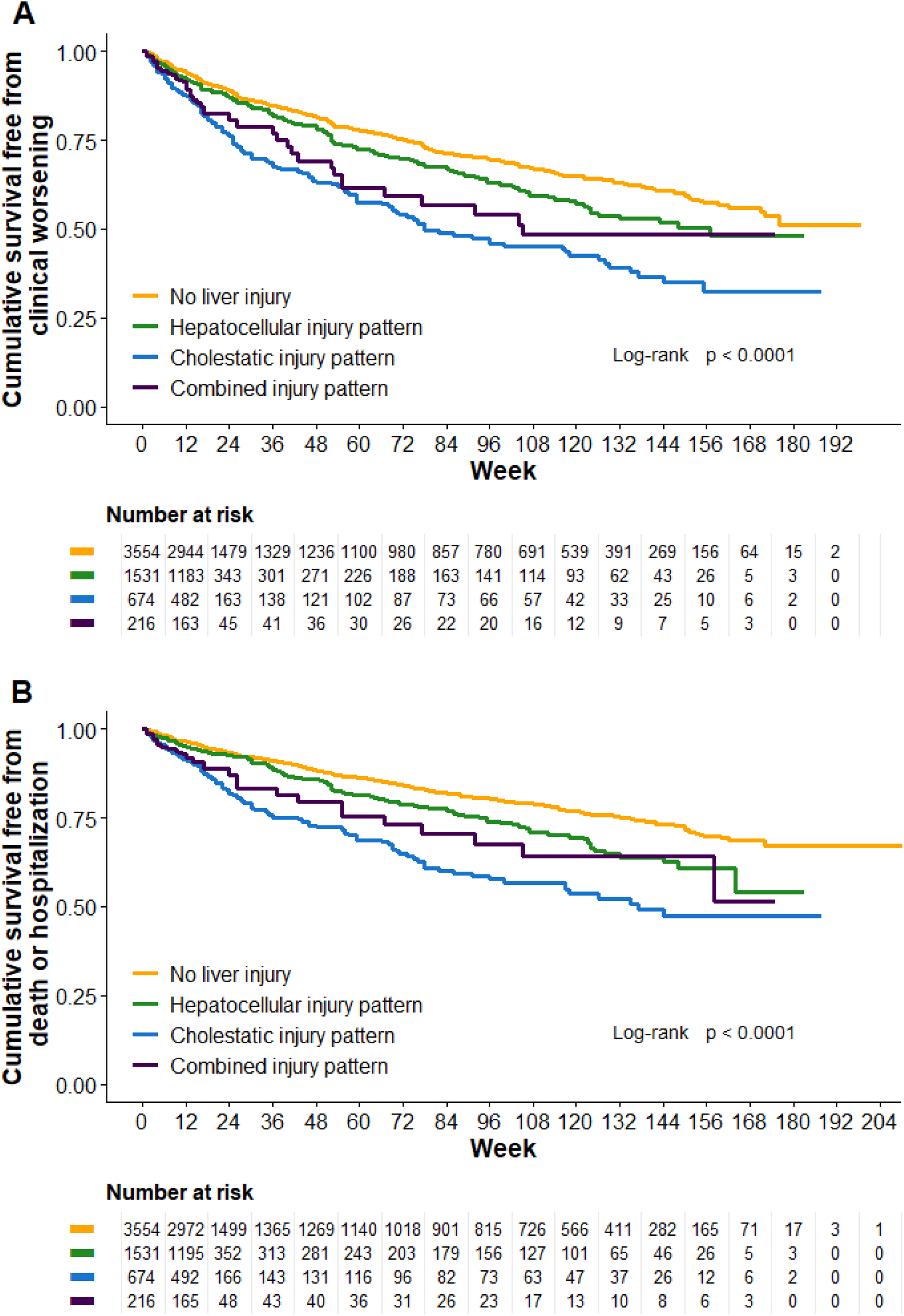
Kapl an-Meie r curv e for time to A) clinic al wors enin g and B) deat h or hosp italiz ation for wors enin g PH.

**Table 3:** Association between baseline liver injury phenotype and clinical worsening events.

| Outcome | Phenotype | Unadjusted HR (95 %CI) | p value | <sup>1</sup>adjusted HR (95 %CI) | p value |
| --- | --- | --- | --- | --- | --- |
| **Entire sample** |  |  |  |  |  |
| Clinical worsening | No liver injury | Ref. |  | Ref. |  |
|  | Hepatocellular injury pattern | 1.36 (1.16, 1.58) | <0.001 | 1.33 (1.14, 1.55) | <0.001 |
|  | Cholestatic injury pattern | 2.19 (1.85, 2.59) | <0.001 | 1.76 (1.48, 2.10) | <0.001 |
|  | Combined injury pattern | 1.85 (1.36, 2.53) | <0.001 | 1.67 (1.23, 2.29) | 0.001 |
| Clinical worsening (Death or hospitalization for worsening PH only) | No liver injury | Ref. |  | Ref. |  |
|  | Hepatocellular injury pattern | 1.53 (1.26, 1.85) | <0.001 | 1.46 (1.20, 1.76) | <0.001 |
|  | Cholestatic injury pattern | 2.55 (2.08, 3.13) | <0.001 | 1.89 (1.53, 2.32) | <0.001 |
|  | Combined injury pattern | 2.16 (1.49, 3.14) | <0.001 | 1.87 (1.28, 2.72) | 0.001 |
| **Subset of studies/subjects with baseline (and if performed, follow up) hemodynamics<sup>2</sup>** |  |  |  |  |  |
| Clinical worsening | No liver injury | Ref. |  | Ref. |  |
|  | Hepatocellular injury pattern | 1.33 (1.13, 1.57) | <0.001 | 1.26 (1.01, 1.56) | 0.041 |
|  | Cholestatic injury pattern | 2.17 (1.81, 2.60) | <0.001 | 1.58 (1.21, 2.07) | <0.001 |
|  | Combined injury pattern | 1.70 (1.20, 2.42) | 0.003 | 1.64 (1.03, 2.62) | 0.038 |
| Clinical worsening (Death or hospitalization for worsening PH only) | No liver injury | Ref. |  | Ref. |  |
|  | Hepatocellular injury pattern | 1.48 (1.20, 1.82) | <0.001 | 1.33 (1.02, 1.74) | 0.035 |
|  | Cholestatic injury pattern | 2.58 (2.08, 3.20) | <0.001 | 1.83 (1.34, 2.52) | <0.001 |
|  | Combined injury pattern | 1.99 (1.31, 3.03) | 0.001 | 1.93 (1.10, 3.37) | 0.022 |

<sup>1</sup>Adjusted for age, sex, body mass index, pulmonary arterial hypertension etiology, baseline six-minute walk distance, and treatment assignment
<sup>2</sup>Additional adjustment for baseline right atrial pressure and cardiac index and (if available) follow-up right atrial pressure and cardiac index as time-varying covariates

The “cholestatic injury pattern” phenotype had a statistically significantly lower change in 6MWD at 12 or 16 weeks while the other phenotypes did not have a significant difference compared to the “no liver injury” phenotype even after adjustment for right atrial pressure and cardiac index (Figure 5). The difference in 6MWD was less than 10 m and of unclear clinical importance. There was no significant association between baseline liver injury phenotype and SF-36 PCS and MCS at 12 or 16 weeks (Figures S2 & S3). Also, there was no significant association between baseline liver injury phenotype and worse WHO functional class at 12 or 16 weeks after adjusting for baseline WHO functional class and other covariates (Table S6).

There was treatment effect heterogeneity by liver injury phenotype (Figure S4). Treatment effects may have been different amongst types of liver injury pattern in terms of time to clinical worsening (but not statistically significantly so), patients with a cholestatic injury pattern had a greater treatment effect in terms of 6MWD and possibly WHO functional class compared to those in the other liver phenotypes.

**Figure 5:**
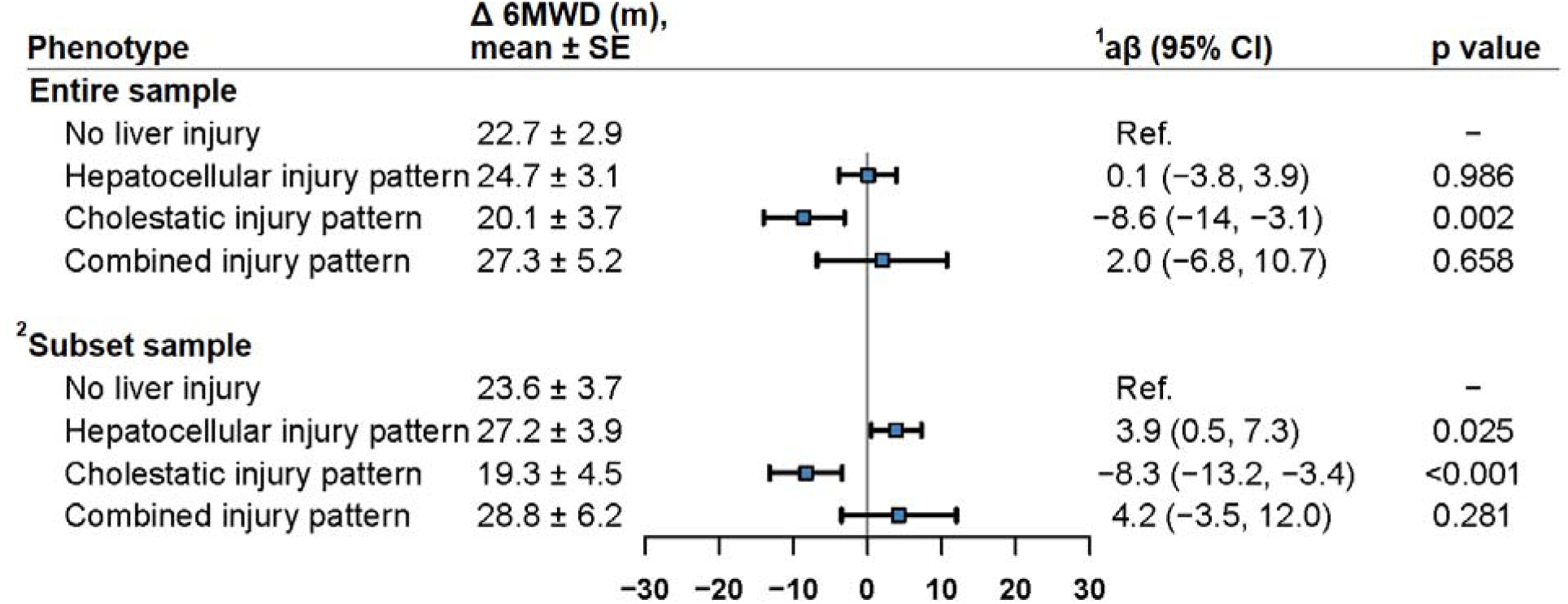
Association between baseline liver phenotype and change in six-minute walk distance at 12/16 weeks. ^1^ Adjusted for age, sex, body mass index, pulmonary arterial hypertension etiology, baseline six-minute walk distance, and treatment assignment ^2^ Subset of studies with baseline hemodynamics with additional adjustment for baseline right atrial pressure and cardiac index and (if available) follow-up right atrial pressure and cardiac index as time-varying covariates

## Discussion

Cardiohepatic syndrome has been increasingly recognized in the setting of both acute and chronic left-sided heart failure, accompanying a greater appreciation of the impact of heart failure on other solid organ function. First identified and defined in congestive heart failure, this syndrome is not well studied in PAH, which is also characterized by increased venous pressure and reduced cardiac output.

In a large dataset of harmonized placebo-controlled RCTs over 20 years, we used five liver tests and unsupervised machine learning to identify four clusters of patients with liver phenotypes which we termed “no liver injury”, “hepatocellular injury pattern”, “cholestatic injury pattern”, and “combined injury pattern”. These clusters (and their characteristics) not only had face validity, but also demonstrated external validity in 1,756 additional patients from two more recent trials left out of the training set. Allocation to experimental treatment significantly reduced the odds of having a worse liver injury phenotype at 12 or 16 weeks.

Those grouped in the liver injury pattern phenotypes had a significantly increased risk of clinical worsening compared to the “no liver injury” phenotype. The “cholestatic injury pattern” phenotype had a significantly lower change in 6MWD at 12 or 16 weeks compared to the “no liver injury” phenotype. Change in WHO functional and SF-36 PCS and MCS at 12 or 16 weeks did not markedly vary by liver injury phenotype. The placebo-adjusted treatment effect on the 6MWD was greater in patients with the “hepatocellular injury pattern” or “cholestatic injury pattern” phenotypes than in patients with the “no liver injury” phenotype, and there were possibly treatment-liver phenotype interactions on time to clinical worsening and WHO functional class, but these were not statistically significant.

Traditionally, abnormalities in liver structure and/or function in congestive heart failure have been attributed to impaired perfusion (i.e., ischemia, usually in the setting of acute critical illness), congestive hepatopathy due to chronically elevated venous pressures, or drug toxicity.^12^ The traditional blood biochemical profile of impaired perfusion includes dramatic increases in transaminases and lactate dehydrogenase, sometimes followed by bilirubin and prothrombin time. The second largest cluster in this study was characterized by variation in (higher) transaminases even if not frankly elevated and despite some of the trials excluding patients with significantly elevated transaminases, leading to a truncated distribution. We have called this the “hepatocellular injury pattern” phenotype, although we do not have liver biopsies to confirm this designation. This group does suggest that cell damage or necrosis (usually seen around the central veins in traditional acute ischemic injury) may occur subclinically in PAH or chronic thromboembolic pulmonary hypertension without critical illness or even abnormally elevated transaminases. Alternatively, variation in transaminases could be a manifestation of drug toxicity, however that would be unlikely in almost one quarter of the patients at baseline in RCTs before starting investigational therapy or placebo. Liver congestion could explain the findings, although right atrial pressure was similar between this group and that of the “no liver injury” cluster. Alternatively, the “hepatocellular injury” phenotype had somewhat lower cardiac index and higher pulmonary vascular resistance compared to the “no liver injury” phenotype, consistent with hypoperfusion as the mechanism for liver injury.

The third cluster (defined by higher total bilirubin and alkaline phosphatase, considered a “cholestatic injury profile”) was most consistent with congestive hepatopathy, usually attributed to increased hepatic venous pressure, decreased hepatic blood flow, and decreased arterial oxygen saturation.^12^ Chronically elevated central venous pressure leads to sinusoidal engorgement and degeneration which can lead to bridging fibrosis, cirrhosis, and even hepatocellular carcinoma. Most congestive hepatopathy is subclinical and commonly overshadowed by the primary cardiac cause.^13^ While standard liver panel tests have not correlated with the degree of fibrosis in congestive hepatopathy, these data are from a limited number of studies focused on Fontan patients rather than PAH or other right heart failure conditions. Supporting venous congestion as a key factor, the “cholestatic injury pattern” cluster had a mean right atrial pressure which was ∼4-6 mm Hg higher than the other groups with lower cardiac index and higher pulmonary vascular resistance (in those with available baseline hemodynamics). This cluster had more men, known to have more severe right ventricular dysfunction in PAH, as well as significantly lower 6MWD and higher BNP levels.^14,15^

The final cluster was comprised of a very small number of patients defined by variation in albumin and a substantial proportion (but still a minority) with a liver diagnosis at randomization.

A few studies have been recently published examining liver function testing in PAH. One study included 407 patients with IPAH at baseline evaluation without known liver disease.^16^ Most (78%) had at least one abnormal liver function test at baseline with hyperbilirubinemia being the most common. Higher total bilirubin was associated with an increased risk of death, after adjustment for confounders including treatment and hemodynamics. Other studies have identified bilirubin as a predictor of outcome in univariate^17,18^ and sometimes multivariate analyses^3,19–21^, suggesting the importance of congestive hepatopathy in PAH. One study showed a possible differential response to therapy in those with higher bilirubin based on the survival of patients.^17^ Lower albumin is the other consistent liver function test linked to a higher risk of death in PAH,^3,21–23^ and may reflect higher severity in many chronic diseases. More recent studies using magnetic resonance imaging have shown that liver stiffness/relaxation times and extracellular volume are higher in the setting of pulmonary hypertension and right heart failure as seen with congestive heart failure.^24^ T1 mapping analysis can assess congestive hepatopathy and liver stiffness and is strongly associated with MR elastography. Hepatic native T1 and extracellular volume fraction of the liver in PAH were associated with an increased risk of death. ^25^ The hepatic native T1 value was not associated with transaminases providing validation of our congestive hepatopathy group (which was not defined by transaminase levels). Other techniques of ultrasound elastography (providing a liver fibrosis index) showed an association with clinical worsening in a very small number of PAH patients.^26^

Short term PAH treatment in these RCTs mitigated the liver injury patterns. Randomization to experimental treatment resulted in a greater proportion of patients moving from a liver disease cluster to the healthy state or from the cholestatic or other injury clusters to a lesser degree of disease (i.e., “hepatocellular injury”) over 3-4 months. During the same time-period, patients receiving placebo were more likely to move from healthy or less severe liver injury clusters to more severe liver injury categories. These results show the impact of PAH therapy (of any type) on liver health (as characterized by liver testing profile) even over the short term. However, we cannot determine the reversibility of the transitions in liver disease from delayed active therapy or the degree of continued improvement on effective treatment as most of the studies were of short duration.

The presence of the “cholestatic injury pattern” or “hepatocellular injury pattern” was associated with an increased risk of clinical worsening after adjustment for potential confounders. While liver phenotype could just reflect hemodynamic severity of disease, this finding persisted despite adjustment for type of pulmonary hypertension and 6MWD at baseline as well as baseline and (when available) follow-up right atrial pressure and cardiac index. The “cholestatic injury pattern” phenotype was also associated with a significantly lower change in 6MWD. Therefore, it is possible that these liver disease profiles could be linked with other processes such as nutrition, inflammation, and metabolic syndrome which may not be adequately addressed by therapies for pulmonary hypertension and could impact on short– and long-term outcomes. Further, these findings may indicate that liver function is an additional predictor of long-term outcomes (i.e. clinical worsening and mortality) to be used in conjunction with hemodynamics. Surprisingly, liver phenotype was not associated with clinically important differences in quality of life over the short term.

There was possible heterogeneity of treatment effect by liver injury phenotype in terms of clinical worsening (p = 0.13) and WHO functional class (p = 0.08), but these were not statistically significant. On the other hand, patients with the “hepatocellular injury pattern” or “cholestatic injury pattern” phenotypes had a significantly larger placebo-corrected treatment effect in terms of the 6MWD compared to patients with “no liver injury”. This suggests that rather than trying to exclude patients with liver injury patterns, studies may want to focus on these patients who appear to gain a greater benefit at least in terms of exercise capacity and possibly functional capacity and clinical worsening. Our findings suggest that liver injury patterns may provide a further insight into response to treatment and the systemic impact of pulmonary vascular disease and RV dysfunction.

This study had several strengths but also some limitations. We derived and then validated our clustering of patients by patterns of liver tests in a large sample of clinical trial participants. The clinical characteristics of the clusters in the validation set were very similar to those of the training set, despite the validation set being distinct in terms of time, background therapy, and other features.^6^ The drugs in these clinical trials are used widely to treat patients with pulmonary hypertension and are approved by regulatory boards based on these studies; our findings should be considered as valid externally as the main results of the therapeutic trials themselves. We focused on efficacy, however there could also be differences in side effect profile, adverse events, and early discontinuations.

Some trials did have exclusion criteria pertinent to liver disease, which may have changed the distribution of livery injury patterns and resulted in spectrum bias. Certain liver tests that might have refined the clusters were not routinely drawn in the sample (GGT, direct bilirubin) and we avoided using international normalized ratio since some of these patients were receiving vitamin K antagonists (and the trials in our validation set did not collect international normalized ratios). However, the completeness of the baseline data (> 99% for all tests except albumin, which was available for > 90%) was one of the strengths of the study. We adjusted for confounders (including baseline and follow-up hemodynamics which were available in a subset of the study sample), however residual or unmeasured confounding is still possible. We had a focused set of hypotheses and therefore did not account for multiple comparisons, which could result in Type I error. This study included 5,975 patients with PAH which is a rare disease so that Type II error is possible (especially in studying effect modification), however this is the largest study of liver function tests in patients with PAH to our knowledge. While we have assumed that subjects in these studies did not participate in more than one study, we cannot be certain of this.

In conclusion, patients with PAH may fall in distinct clusters based on patterns of liver tests. These clusters were reproducible in clinical trials left out of their derivation. Patients with cholestatic injury or hepatocellular injury patterns have an increased risk of clinical worsening. While treatment effects of PAH therapy on clinical worsening do not appear to differ based on these liver phenotypes, there may have been a larger treatment effect in patients with liver disease compared to healthy status. Future studies need to pursue a better understanding of the contribution of liver disease to PAH and the effect of therapies on outcomes.

## Data Availability

Data is not broadly available to the public nor open-source, select data may be available to reviewers upon request

## Sources of Funding

Jacqueline Scott has received consulting fees for the preparation of this manuscript from University of Pennsylvania; Robyn McClelland has received FTE support via subcontract to UPenn for this manuscript; Steven Kawut has received funding from the NIH and Cardiovascular Medical Research and Education Fund; Jasleen Minhas has received the ATS Early Career Investigator award in PVD; Nadine has received grant funding from NIH/NHLBI K23HL141584

## Disclosures

Jacqueline Scott is an employee of Werfen, was previously a paid research fellow with the U.S. Food and Drug Administration, received honoraria for a lecture at Stanford in 2020, has received consulting fees for the preparation of this manuscript from University of Pennsylvania; Robyn McClelland has received grants and contracts from the NIH; Steven Kawut has received consulting fees from Janssen, Regeneron, and Morphic; Steven Kawut has received payment or honoraria from Janssen, Accredo, Actelion, Aerovate, Bayer, Inari Medical, Merck, United Therapeutics, Janssen, Liquidia, and Pfizer for lectures, presentations, speakers bureaus, manuscript writing or educational events; Steven Kawut has received support for attending meetings and/or travel from Aerovate; Steven Kawut has participated on the following Data Safety Monitoring Boards or Advisory Boards: DSMB Chair, United Therapeutics, Acceleron, Ad Board, United Therapeutics, Study section, Vivus, Ad Board Aerovate, Ad Board, Steering Committee, Proteo Biotech; Steven Kawut is a paid member of the European Respiratory Journal Advisory Board; Steven Kawut has stock options with Verve Therapeutics; Ethan Weinberg has received consulting fees from BioVie and PharmaIN; Ethan Weinberg has received payment from Institute for Medical and Nursing Education for lectures or other related events; Ethan Weinberg is on the Data Safety Monitoring or Advisory Board for Mallinckrodt; Harold Pavelsky is on the Data Safety Monitoring Board for Ralinepag in PAH – being developed by United Therapeutics; Jasleen Minhas has received the ATS Early Career Investigator award in PVD; Corey Ventetuolo has received NHLBI grant funding and grants from Altavant Sciences, clinical trial participation; Corey Ventetuolo has received consulting fees from Regeneron, Merck, Janssen; Corey Ventetuolo has received payments or honoraria for lectures or other related events from Gather-ED CME; Corey Ventetuolo has received support for attending meetings or travel from the American Thoracic Society ICC; Corey Ventetuolo is on the Data Safety Monitoring Board for U01 Saturn and Altavant Sciences prior advisory board and Acceleron/Merck prior advisory board; All other authors have no disclosures for competing interests within the past 36 months.

## Notes

### Author Declarations

All trials were performed under the supervision of human ethics committees, and all patients provided informed consent to participate in the trials. Harmonization and analysis of these data were exempted from review by the University of Pennsylvania institutional review board.

